# Health Heatmap of India: An Open Data Platform

**DOI:** 10.1101/2020.09.18.20197699

**Authors:** Akshay S Dinesh, Varnita Mathur, B. R. Ansil, Vijay Chandru, Ravi Chellam, Abi Tamim Vanak, Uma Ramakrishnan, Prabhakar Rajagopal

**Author notes:** This manuscript will appear in the Journal of Indian Institute of Science, Vol 100-4, October 2020.

## Abstract

Health Heatmap of India is an open data platform built for bringing together data from diverse sources and facilitating visualization, analysis, and insight building from such data. In this paper, we describe the context and need for such an open data platform and describe the technical aspects of building it. The beta site of the portal is available at http://healthheatmapindia.org

## Introduction and importance of open data in Health

Open data is a movement that is gaining momentum all over the world. It not only advances science and scientific communications as open science^1^, but it also has the power of transforming societies and the process of informed decision making. The availability of open data in different scientific and social spheres contributes to enabling and establishing transparency and accountability, essential pillars of a democratic society. It supports informed discussions and debates among citizens, scientists and governments and drives evidence-driven policy planning^2^. Nobel laureate Elinor Ostrom viewed open data as a public good to be shared with citizens and the community as commons^3^. Open data is driven by four foundational principles - Findability, Accessibility, Interoperability, and Reusability, (FAIR) that should guide all producers and publishers of data^4^. The large scale availability of open data will help build synergies and collaborations across institutions and disciplines.

In the domain of health, the value of open data for research as well as for public health delivery systems, is significantly enhanced by combining datasets on public health with other environmental, social and geographical data^5^. Effectively combining environmental and land use datasets with health data has helped in building risk landscapes for infectious diseases and for predicting the progress and spread of diseases^6^. The ability to plan and provide health infrastructure for communities can be helped by a spatial study of health infrastructure and the social and economic distribution of communities and habitations^7^. The NITI Ayog of the Government of India has used open data in health to create a Health Atlas and along with other indicators, to identify Aspirational Districts^8^. The One Health approach to human health and well-being, adopted by the Government of India, involves human health, environmental health and animal health components and requires multi-sectoral transdisciplinary collaborations to achieve better public health outcomes. Assembling and combining open data across domains is absolutely vital for a One Health approach to human health ^9, 10^.

The current COVID-19 pandemic has exposed the vulnerability of human societies to zoonotic diseases. The pandemic requires careful monitoring and near real time availability of open data for governments, societies and citizens to manage and control the disease. Many such government, research and volunteer driven initiatives have sprung up to aggregate and provide open data. Some like the Johns Hopkins University, Coronavirus resource center https://coronavirus.jhu.edu/map.html and volunteer-driven crowd sourced efforts https://www.covid19india.org/ are effective and widely used. They provide regularly updated and near real time dashboards on a spatial platform on the status and spread of the pandemic. These initiatives need to be sustained in the post pandemic world providing reliable open data on health and diseases with a wide range of related interdisciplinary variables.^11^

The Health Heatmap (HHM) initiative which is a part of the preparatory phase project of the National Mission for Biodiversity and Human Well-Being^12^ supported by the Office of the Principal Scientific Adviser to the Government of India, focuses on developing an open data platform for aggregating data. The HHM will integrate open data on health with the documentation of India’s biodiversity, traditional knowledge and cultural practices related to medicinal plants and their conservation.The HHM is intended to be an important resource for the One Health initiative, integrating human health data with environmental and animal health. This will aid in better understanding of spatial and temporal patterns of prevalent infectious diseases specifically zoonotic and vector-borne diseases.

It is urgently necessary to survey existing data aggregation and curation platforms to develop a HHM platform with open data built on FAIR principles. The value of a health heatmap in digital health research and policy analysis needs to be emphasized. Underlying the trend visualizations that the heatmaps so effectively convey, are quantitative representations or models that can be associated with research agendas in digital health that will inform policy. Systems and game theoretic models in understanding public health policy levers (See Fig 1) or agent-based simulation models of epidemic risks are evident examples.^13^ In summary, a mature health heatmap can enable multiple research agendas in analytics that can direct public interventions and advise policy. It is crucial to recognise that the first building block is the assembly and availability of open data conforming to the FAIR principles that will enable subsequent benefits in research, policy, public health management as well as an informed and enlightened citizenry.

**Figure 1:**
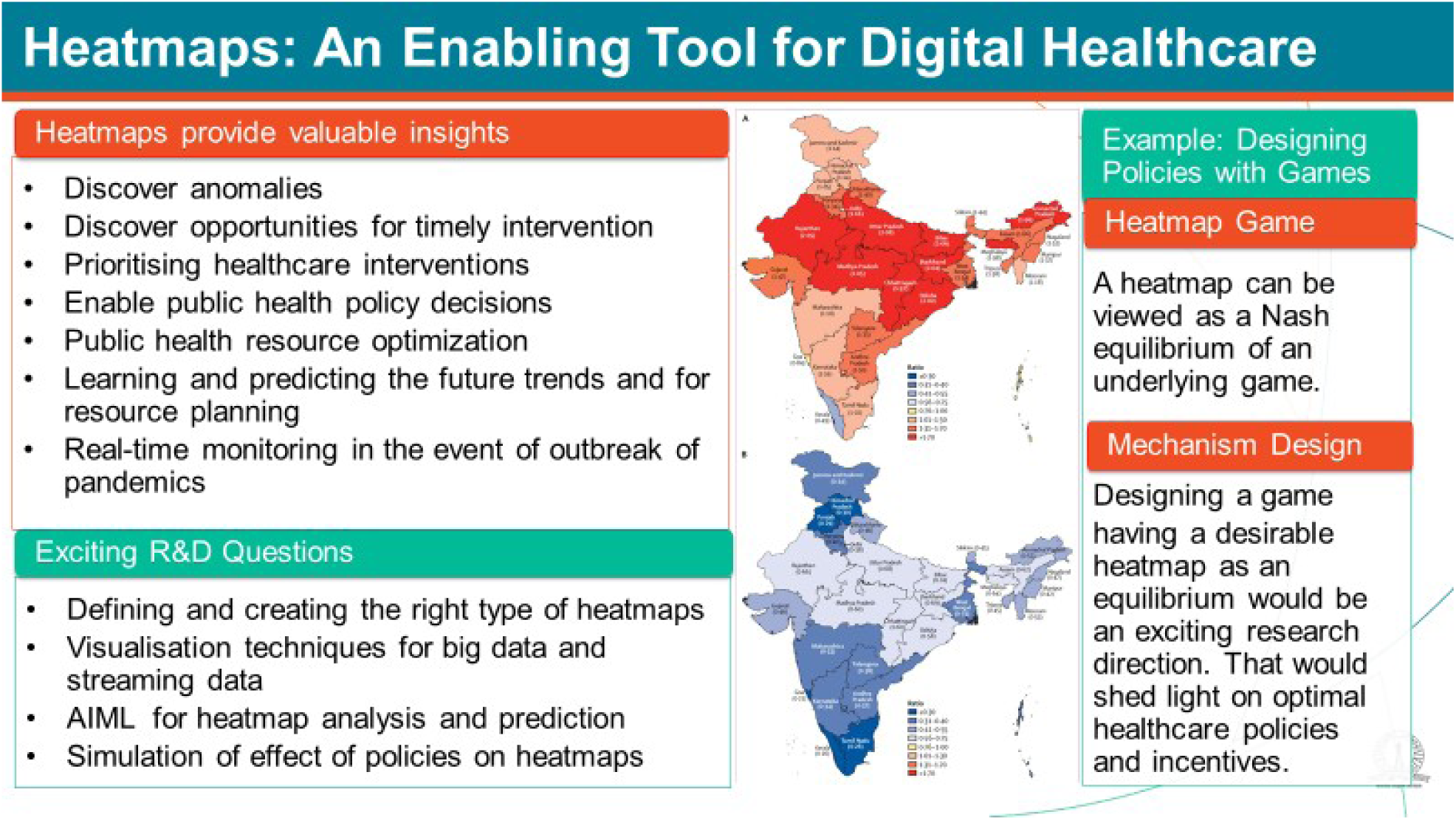
A schematic of health heatmaps^14^

## Heatmaps: An Enabling Tool for Digital Healthcare

Health heatmaps created with extensive data are valuable in many ways. They could be used to detect anomalies in the healthcare landscape. They could be used to discover opportunities for timely intervention and also to prioritise healthcare interventions. They provide insights for public health resource optimization and for learning and predicting the future trends for better resource planning. When produced from continuously streaming data, heatmaps provide the basis for real-time monitoring and planning in the event of a disease outbreak. Studying heatmaps can provide critical inputs for forming better public health policies. In other words, heatmaps provide an enabling tool for strategic, tactical, and operational decision making.

There are many exciting R&D questions that healthcare heatmaps throw up. Defining and creating the right type of heatmaps is a critical need. Visualisation techniques for big data and streaming data provide interesting challenges. Novel AI/ML techniques are needed for heatmap analysis and prediction. Use of efficient simulation and gaming approaches for rigorously evaluating the effect of policies on heatmaps is another interesting direction.

Figure 1 is indicative of several applications that can be built on top of the foundation of a health heatmap. The gist of this paper describes an attempt to create the first box that instantiates how heatmaps provide valuable insights.

While building out the R&D directions indicated in Figure 1 will remain an agenda for future collaborative work, we provide a glimpse of some of these below.

Analytical methods for building simulation models that could guide policy are sometimes called phenomenological or mechanistic models. These models take as inputs various hypotheses and output descriptions of the health systems.

There is also a data driven methodology for modeling that takes data as input and outputs a summary of data. Such models would be called statistical models or data-analytical. A third approach would be models that take data as input and will output predictions. Such models come under the classification of machine learning or AI/ML models.

Another direction depicted in figure 1 may be thought of a dynamical systems view of the agencies of a health system. Typical systems properties such as robustness or resilience can be studied within such a framework.

The heatmap that shows the state of healthcare (for any disease or pandemic) could be viewed as a Nash equilibrium of a game that captures the complex dynamics of the disease, Government policies, policy implementation protocols, and individual compliance. One can ask the following research questions:

- How can we create the heat map of a resilient healthcare ecosystem?
- What is the game that has the above (desirable) heatmap as a Nash equilibrium? In game theory parlance, this is a mechanism design problem^15^.

Indeed a research agenda that encompasses phenomenological to predictive to resilient health systems will be the next stage that builds on the Health Heatmap of India.

## Survey and review of global platforms on health data

Public health data is distributed on multiple platforms across the world, each with different objectives, target groups and functions. (a) There are many country and state level platforms that aggregate and publish data and statistics on public health. These are normally government statistics offices that gather data from various departments and publish them as open data. For example, the Office for National Statistics of the United Kingdom publishes data on various aspects related to the health of the human population. The European Union assembles and publishes data on the health of human populations of its member states as an annual report.^16^ The Government of India aggregates health data from all the states and publishes them as open data on the national portal, data.gov.in. These are meant to provide researchers, planners and citizens in general, information on public health. (b) There are specific thematic platforms that gather and provide information for monitoring and management of public health. For example, the centres for disease control in various countries gather data on infectious and non-infectious diseases that feed into relevant departments of the state and some of this information is provided as open data. (c) At the global level, international and multinational agencies like the World Bank and the World Health Organization maintain an extensive data repository called the Global Health Observatory (GHO) on various health indicators for countries and regions around the world. This is helpful for global policies on human health to advise countries, suggest policies and focus global attention on health issues in all the countries of the world. Projects like Health Intelligence (http://publichealthintelligence.org/) aggregate public health data across the world and add rich visualizations of the data to provide interesting insights for managing and improving public health.

In this section, we explore the functions, features, and data organization of three important global open data platforms - the Global Health Observatory, the World Bank Open Data portal and the open source District Health Information System (DHIS 2).

WHO’s Global Health Observatory (GHO) is an open online public health observatory^17^ setup with the purpose of “making health data easier to find and use for specialists such as statisticians, epidemiologists, economists and public health researchers as well as anyone with an interest in global health.” ^18^ It is built over the extensive data collected by WHO from its 194 member countries on more than 1000 indicators of priority health topics including mortality and burden of diseases, the Sustainable Development Goals, non communicable diseases and risk factors, epidemic-prone diseases, health systems, environmental health, violence and injuries and equity.

WHO sources its data from government’s registration records of births and deaths, health systems, surveys and censuses, research projects and databases maintained by other organizations. To allow comparison across countries and across time, datasets are normalized using appropriate methodologies. Hence, they are not always the same as official national estimates. To give users of the data the necessary context to interpret these correctly, an Indicator Metadata Registry is maintained. The indicator metadata contains the protocols for collection and sourcing of the data, the type of the indicator and the aggregation method.

The Global Health Observatory portal has been redesigned in the beginning of 2020 making it easier for navigating the numerous health indicators that are tracked. This is achieved by organizing indicators into themes which are further divided into topics. There are landing pages for each theme and topic which give a high level overview. On the same page a user is able to dig further in and access and explore more specific data^19^. These features and redesign have helped greater accessibility and use of the data.

The GHO operates at the scale of nations providing aggregates at the national level to give an overall status of the global health of the human population. However, for policy and action at the local community level more detailed information is required. Such global platforms provide the model, framework and inspiration to build information systems that can hold data at the granularity of the state, district, and local community level.

The World Bank has also been systematically collecting, generating, and publishing open data. They work with member countries, including by investing in statistical activities and creating standards and frameworks for data collection. The World Bank focuses their effort on global development data, but this includes a large number of health indicators along with other social and economic indicators.

Recognizing the importance of open data in economic development, the World Bank started releasing their datasets under Creative Commons Attribution 4.0 International License in the year 2010. They have also developed an analysis and visualization tool called DataBank which allows users to craft custom queries and generate and share charts, maps, and tables.

Additionally they maintain a data catalog which allows searches through the datasets, and a microdata library which is a catalog of highly granular datasets from various surveys and studies.

The District Health Information System (DHIS) is a comprehensive open source health information platform that was initiated in 2006 and has been steadily growing to over 90 installations today. It is an efficient and low budget platform specifically suited for health data collection, monitoring and reporting from distributed health centres, data management and analysis. It is easy to install by an IT systems team and there are a few service providers who undertake the installation, configuration and support of the DHIS system.

The data model is flexible and supports defined data elements, data values and hierarchy of organizational units. However once designed, alterations, changes and additions to the data model is difficult. Thus it works very well in a structured health delivery system at various levels of the Government. A configurable and interactive dashboard with a spatial map component is an attractive feature of this platform.

It is extensively used by many low and middle income countries of Africa and Asia and is extensively piloted and used by 22 states of the 28 states and 8 union territories of India. However its efficacy, extent of use and integration with national health statistics in India have not yet been ascertained. WHO packages and toolkits have been incorporated in 30 DHIS deployments across the world.

## Open data on health in India

The Government of India, in 2012, formulated the National Data Sharing and Accessibility Policy (NDSAP) which draws inspiration from the Right-To-Information (RTI) Act, 2005 and the United Nations Declaration on Environment and Development (Rio, 1992). In it, the role of open data in increasing public awareness and participation in decision making processes has been explicitly recognized. There are various Indian sources of open health data which contribute to public health research, policy making, awareness, and overall human development.

The single largest source of health data in India is the National Health Mission (NHM). Through the Health Management Information System (HMIS) portal they aggregate data from all over India and publish them in periodic reports. These include data on more than 500 indicators available at district and sub-district level, starting from 2008-09.

NHM also releases publications like the health and family welfare statistics which combines and compares various data sources like the Population Census, Sample Registration System, National Family Health Survey, District Level Household Survey, and others.

Similarly, the Central Bureau of Health Intelligence under Directorate General of Health Services releases an annual “National Health Profile” combining data from central ministries/departments, state health authorities, and autonomous organizations / other agencies. But the tables in these publications are aggregated at the state level.

Data at higher granularity (district or sub-district) is often available on the website of each of these sources. We list some of the major sources in table 1 while describing their importance.

**Table 1:** Major sources of health data in India

| Source | Description | Periodicity |
| --- | --- | --- |
| Civil & Sample registration system (SRS) | Details about births and deaths that make it possible to extrapolate values | Continuously collected and reported periodically |
|  | between censuses, and details on birth rates and death rates. |  |
| Health Management Information System (HMIS) Standard & Analytical Reports | Web-based portal set up by the National Health Mission that collects data from district and state authorities on various health indicators | Monthly reports generated from continuously collected data |
| National Family Health Survey (NFHS) | This large-scale, multi-round survey is conducted in a representative sample of households throughout India and contains questions related to maternal health, child health, nutrition, awareness about diseases, immunization, etc which gives rise to more than 100 indicators. | The first survey was conducted in 1992-93, and the next rounds have been in 1998-99, 2005-06, 2015-16, and 2018-19. |
| District Level Household and Facility Survey (DLHS) | Closely related to NFHS, DLHS also generates data about reproductive and child health. | This survey was done in 1998-99, 2002-04, 2007-08, and 2012-13. |
| Annual Health Survey (AHS) | Now discontinued, this survey included data from 8 Empowered Action Group states and Assam on various parameters. | Annually, from 2010 to 2013. |
| Integrated Disease Surveillance Program (IDSP) | Under the National Center for Disease Control, IDSP has established a state based surveillance system for epidemic prone diseases. | Weekly reports of disease outbreaks are published on their website, starting from mid-2009. |
| Census | The Population Census includes demographic data related to age, gender, social group (SC/ST, OBC), urban-rural divide, etc that form the denominator to various health indicators. | Every 10 years |
| Livestock census | The Department of Animal Husbandry and Dairying does a periodic census of livestock and poultry in the country and makes this data available. | Every 5 years |

A lot of data from sources like these eventually make their way to the Open Government Data (OGD) Platform of India (data.gov.in) which was setup in 2012 as a result of the National Data Sharing and Accessibility Policy (NDSAP).

Newer platforms like Integrated Health Information Platform, Ayushman Bharath Yojana, and National Health Stack are slated to capture and make available machine readable data of a larger volume and velocity, although presently they cannot be considered as sources of open data.

There are various other sources of data including those generated by public health researchers through studies and those contributed by citizens through citizen science platforms that contribute to health data. We take a look at how such private efforts compare with national programs in the case study of data on infectious diseases below.

## A qualitative comparison of IDSP and ProMED reporting platforms

ProMED is an initiative of the International Society for Infectious Diseases (ISID) for reporting of infectious disease outbreaks affecting humans, animals and plants across the globe. Reports in this portal are produced and commented upon by domain experts from a variety of fields related to public health including epidemiology, virology, veterinary and plant diseases. These reports are an important source of information for clinicians and researchers by providing timely reporting on emerging pathogens and their vectors at a global scale.

The Integrated Disease Surveillance Program (IDSP) is hosted by National Centre for Disease Control (NCDC), Govt of India with an objective of decentralized, technology-supported disease surveillance system for epidemic-prone diseases. One of the important components of IDSP is inter-sectoral coordination for zoonotic diseases. The reporting in this portal is done by health workers, clinicians and laboratory staff on a weekly basis. This data is analysed by respective district surveillance units in real-time and investigated by the rapid response teams.

Both IDSP (https://idsp.nic.in) and ProMED (https://promedmail.org) are surveillance platforms with the objective of reporting infections in real-time towards assisting prompt detection and mitigation responses for disease outbreaks. Though there is a difference in geographic coverage in reporting, the information generated is mutually complementary and can act as an important resource for clinicians, veterinarians, healthcare workers, researchers, government officials and the general public.

There are 22 listed diseases/categories which are generally reported by the IDSP platform, which include infectious diseases, food borne illnesses, snake bites and dog bites. However, few categories in this list such as ‘state specific diseases’ and ‘unusual syndromes’ provide freedom to the professionals to report new infections and symptoms of unknown origin. For example, emerging infections such as Nipah, Brucellosis and Anthrax are not included in the list but still these are getting reported because of these general categories. In the case of ProMED, the main focus is on emerging and re-emerging infections of humans, wildlife, livestock and plants. This comprises a wide variety of infections including viral (Crimean-Congo haemorrhagic fever (CCHF), Japanese encephalitis, Kyasanur forest disease etc.), bacterial (Scrub typhus, Anthrax, Brucellosis etc.) and protozoan infections (Malaria and Leishmaniasis). However, regular recurring tropical vector-borne infections (e.g. Chikungunya and Dengue) are not reported on this platform.

We accessed the incidence of zoonotic and vector-borne diseases from IDSP and ProMED for 2019. The data obtained from IDSP was specifically filtered for known zoonotic as well as vector borne diseases reported in 2019. Reports of infections outside this category, general symptoms, multiple infections without confirmation and infections of unknown origin were omitted during the filtering. In the ProMED portal, a search was conducted with a specific filtering keyword “India” for 2019. From these, reports of zoonotic and vector-borne infections were manually collected and curated. Data from both the sources were tabulated and compared for attributes such as type of diseases, geographic coverage, and number of infections reported per disease.

We noticed a wide variation in the level and quality of reporting from various states. Zoonotic and vector-borne diseases data for many states and union territories are poorly represented in both platforms (Figure 2). Besides, states such as Mizoram and union territories such as Andaman & Nicobar Islands and Lakshadweep were not represented in both the platforms. This high variability in the number of reports suggests that there are inherent biases as well as non-uniformity in reporting across states. This needs to be further investigated to understand confounding factors creating these reporting biases. However, IDSP has more geographical coverage and number reports for zoonotic and vector-borne diseases in 2019. The same can be visualized in the map below (Figure 3) which has been created by collecting geo-coordinates for each district from Google Maps API^20^ and plotted using QGIS3.10^21^. We have attempted a similar exercise with ProMED data to achieve a comparative understanding of the effectiveness of both these platforms in their geographical coverage of reporting diseases from India (Figure 2). Since the ProMED data are often consolidated state-wise, and not at a finer granularity, we were unable to perform direct comparisons with the data reported on IDSP.

**Figure 2:**
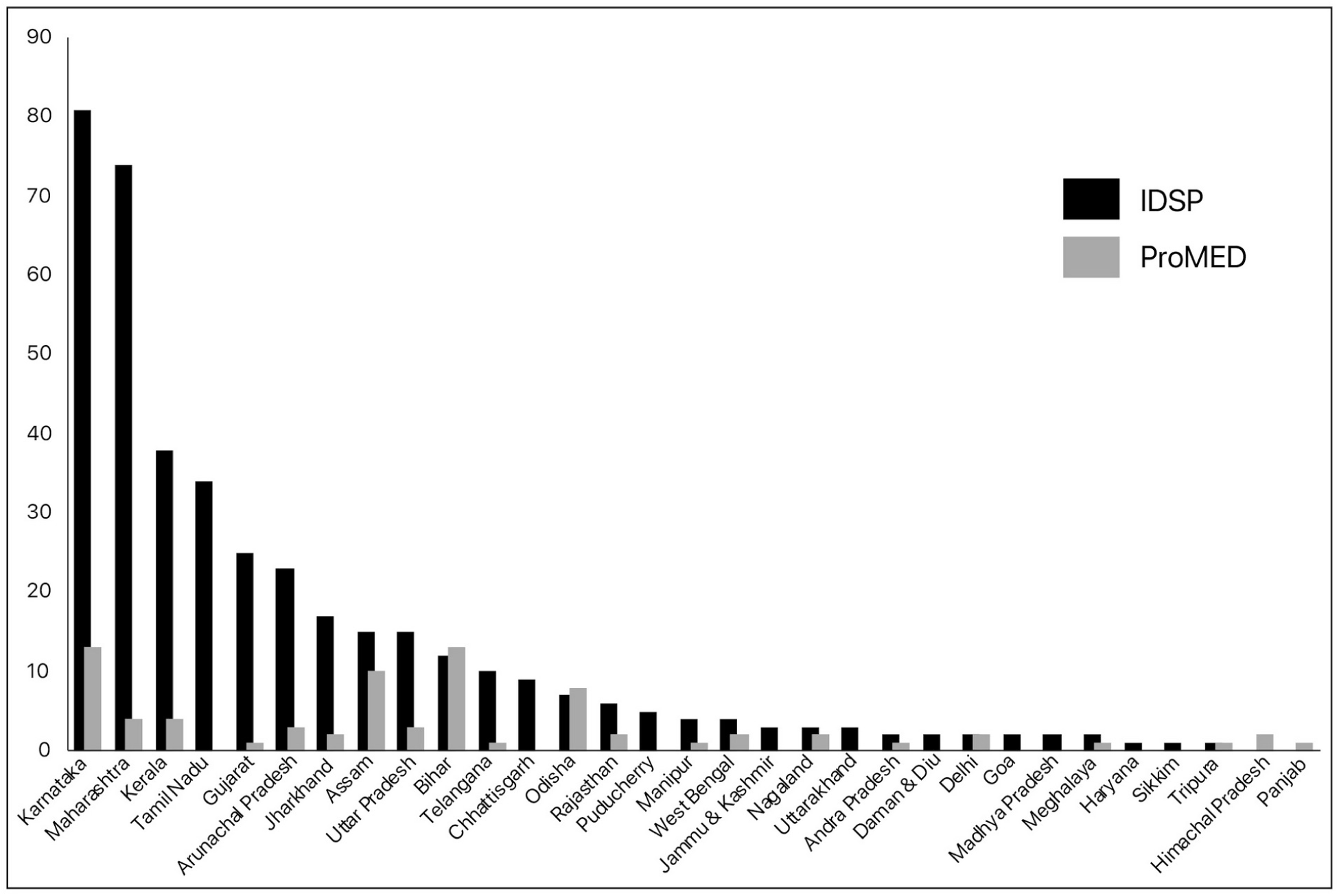
State-wise number of reports of zoonotic and vector-borne diseases reported in IDSP and ProMED platforms during 2019.

**Figure 3:**
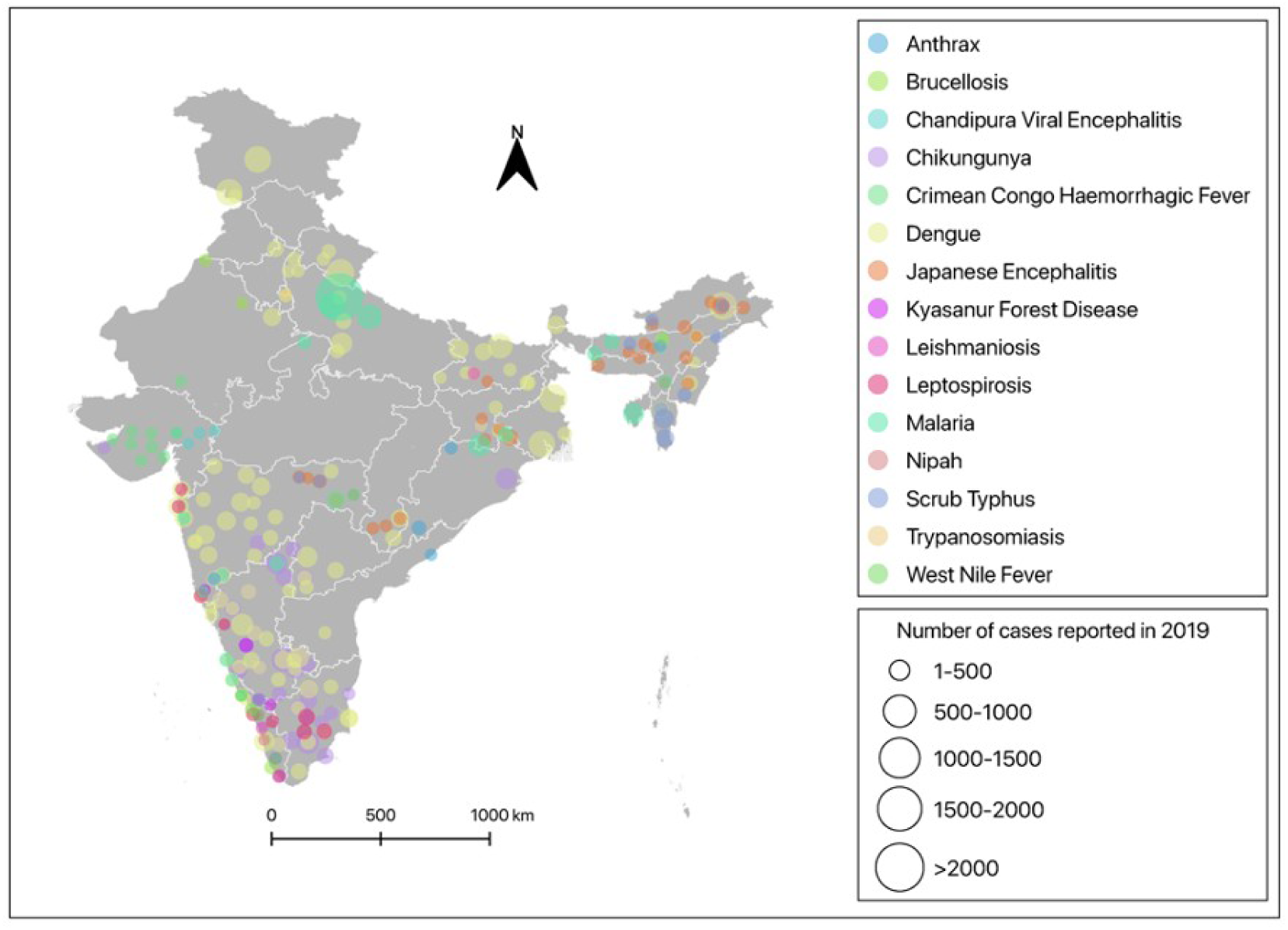
Map showing the geographic locations of zoonotic and vector-borne diseases reported in IDSP during 2019

In both platforms, district names are reported as outbreak location and occasionally village name, block and ward are mentioned. In the case of district-level locations, it is difficult to trace accurate geographic coordinates and this hinders the future use of data for understanding environmental correlates of disease outbreaks. However, the user-friendly interface, representation of reports on an interactive map, and availability of email subscription to the public are unique features of ProMED. Table 2 summarises our qualitative comparison of different attributes of these platforms.

**Table 2:** Comparison between IDSP and ProMED

|  | **IDSP** | **ProMED** |
| --- | --- | --- |
| Hosted by | NCDC, Govt of India | ISID |
| Initiative type | Surveillance | Surveillance |
| Geographical area | India | Across the globe |
| Reporting time unit | Weekly-throughout the year | Based on cases reported in the media |
| Reporting methods | Direct-based on incidence | Indirect-based on news or media coverage |
| Reporting personnel | Health workers, clinicians and laboratory staff | Subject matter experts (SMEs) |
| Review process/analysis | District surveillance units | ProMED staff and moderators |
| Disease coverage | Epidemic prone diseases of humans | Emerging and re-emerging infections of humans, animals and plants |
| Geographic unit | District level | Varying. Sometimes village level information |
| User interface | Not interactive | Interactive and user friendly |

\*These comparisons are based on 2019 data reported in IDSP and ProMED

**Table 3:** The health indicator structure on the HHM platform.

| Theme | Subthemes | Number of indicators available as of now |
| --- | --- | --- |
| Infectious Diseases | • Air-borne diseases<br>• Water borne diseases | 8 |
| Child Health | • Nutrition<br>• Mortality<br>• Sexual & Reproductive health<br>• Vaccination | 42 |
| Adult Health | • Awareness<br>• Lifestyle hazards<br>• Nutrition | 5 |
| Maternal & Reproductive Health | • Antenatal care<br>• Delivery services<br>• Postnatal care<br>• Family planning<br>• Sexual & Reproductive health<br>• Mortality<br>• Nutrition | 44 |
| Non-communicable Diseases | • Asthma<br>• Cancer<br>• Diabetes<br>• Hypertension<br>• Heart disease<br>• Obesity<br>• Thyroid disorder | 17 |
| Health Infrastructure | • Health services<br>• Health surveillance<br>• Essential medicines<br>• Universal health coverage | 18 |
| Demographics | • Birth and death rates<br>• Demographics<br>• Gender-preference | 10 |
| Social Development | • Development Indicators | 18 |
|  | • Education<br>• Women's empowerment |  |

## Disease Surveillance and Epidemiological Models

As noted above, both IDSP and Promed are data banks which have been created and maintained to support disease surveillance required for public health research as well as policy and intervention. Classical epidemiological models built on these data using biostatistics and compartment models have been used over the years to depict and predict the dynamics of infectious disease spread in populations and have advised public health authorities with strategies for coping and suppressing epidemics when they break out. These are sometimes called phenomenological mechanistic models since they are based on principles worked out by the scientists on the mechanisms of the spread of disease.

Traditional surveillance would typically involve a combination of screening and rigorous testing methodologies with specimen samples collected from the population. Syndromic surveillance on the other hand observes clinical features that are discernable before diagnosis is confirmed or activities prompted by the onset of symptoms as an alert of changes in disease activity.

Occasionally, the mechanistic models can be abstracted into a logical framework of a rule based or axiomatic representation. Rules of logical inference using classical deductive reasoning or the modern inductive and abductive reasoning methodologies popularized in causality research in artificial intelligence (AI) can be useful in generating explanations for epidemiological outcomes. In a communication that appears in this issue, using this methodology, an anecdotal explanation is described for the zoonotic transmission of KFD in Karnataka^22^.

A third emerging approach to surveillance and forecasting is in the use of data science to improve the predictive accuracy of the classical epidemiological models. We call these the statistical epidemiological models which of course are largely dependent on data. The recent widespread use of these modeling efforts in the context of the raging pandemic of COVID-19 are well known and are also described in several articles in this issue of the journal. The article by Adiga et al.,^23^ in this issue presents a modern overview of the use of mathematical models in the context of a pandemic like COVID-19. The quality and predictive power of the models built will largely depend on the quality of data repositories that we build and maintain for current and future disease burdens of the country.

In the midst of a global pandemic it has been clear that due to the paucity of good data, epidemiological models have been effective largely for counterfactual analysis but not for forecasting in a reliable way. Perhaps we can be better prepared for the next one with the right data frameworks for disease surveillance.

## Framework for health data warehousing and management

The Health Heatmap is conceived as an aggregation platform that integrates public health data from a variety of sources. Data in health is highly heterogeneous, available in various formats and of variable frequency and quality. The granularity of data varies between each source, and is often associated with different attributes. A unified health indicator - geo entity framework and data model allows disaggregation of the incoming data so that it can be re-combined in various ways depending upon the questions being asked of the data.

Each data element is referenced on a spatial dimension or geo entity of a state, district, tehsil (sub-district), village or a latitude/longitude location. This allows the flexibility of aggregating data of different granularity, scale and extent onto a common framework. The spatial reference allows health data to be easily combined with other environmental, economic and social data and visualize the health data along with other dimensions on a map visualization engine. This is especially helpful in tracking and tracing infectious diseases and exploring patterns in environmentally or socially mediated diseases.

Each data element is associated with a health indicator. These health indicators are grouped into an hierarchical ontology. This grouping allows intuitive navigation, search and access of the data as well as facilitate building combinations of indicators and ranking them to provide a comprehensive view of public health status across geo entities. These indicators are also mapped to the Sustainable Development Goals (SDGs) that allow cross comparison in relation to global development efforts. The indicator framework also provides the flexibility to add indicators into the ontology structure depending upon the need as well as availability of data. The health indicator framework is most commonly used in global platforms like the World Health Observatory (WHO) and the DataBank of the World Bank.

Each data element is annotated with metadata explaining the indicator as well as the source, protocols of data collection, the computation and aggregation rules and the positive or negative nature of the indicator with regard to overall health outcomes. Each data element with all its dimensions are individually stored which allows flexibility in search, query, and aggregation depending upon the use cases as well as user requirements.

## An extensible data model for health data

The HHM Platform is built with a set of microservices. Figure 4 gives a high level overview of the components of the health heatmap portal.

**Figure 4:**
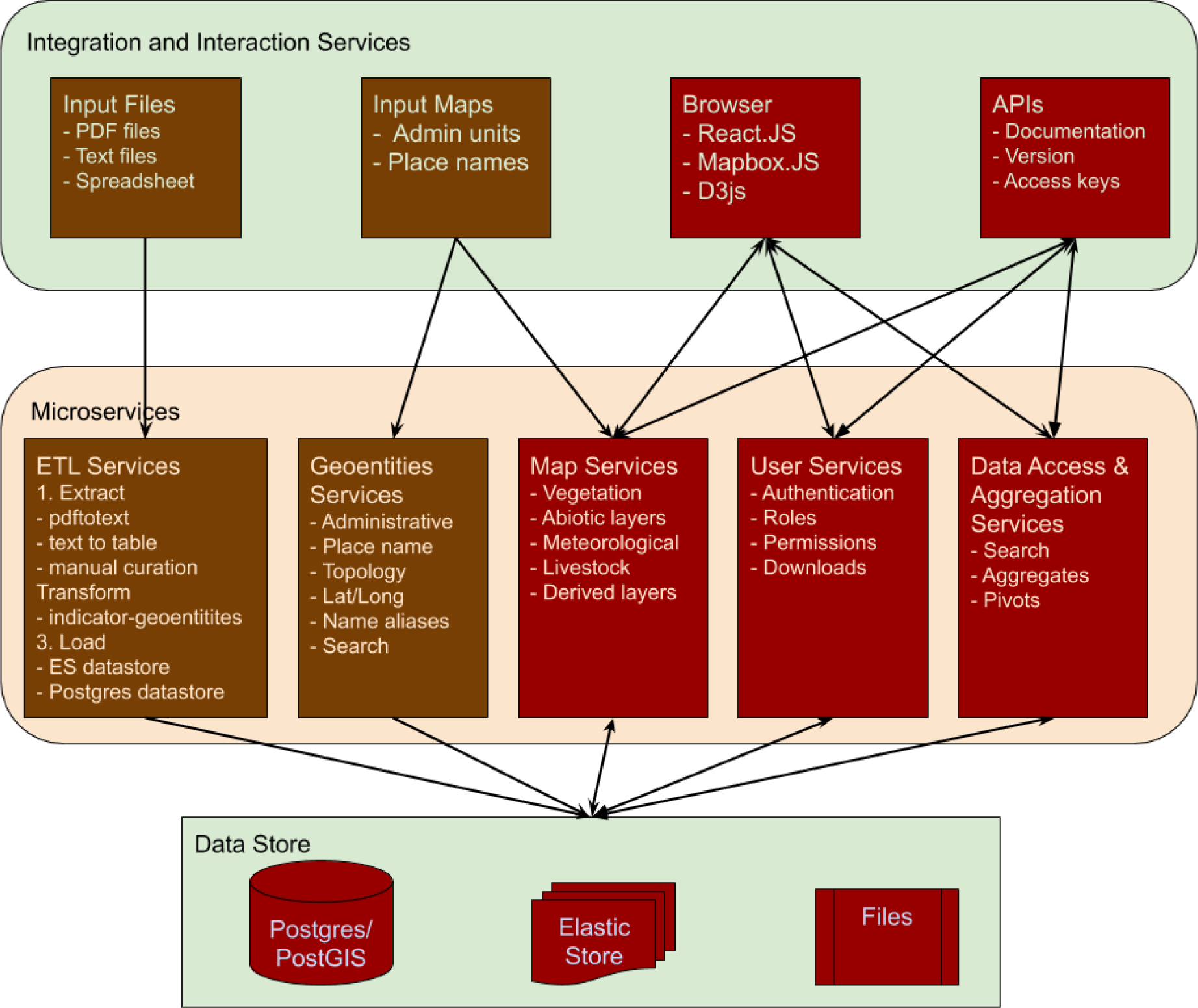
HHM Architecture.

### Data Pipeline

Open data from multiple sources listed above are in various formats as tables, text and pdfs. The data pipeline is essentially an ETL (Extract, Transform and Load) service which enables differently formatted data to be warehoused in a common structure.

The extract function is a semi automated process where data is extracted from pdf and text through an automated process and manually inspected and curated. Metadata is created and associated with the datasets. All stages of the extraction process are archived as source files, files generated from PDFs, curated files, and metadata files. This allows an audit of the extraction process and the ability to trace data back to source.

The transform process creates a data point for each extracted value, associating it with an indicator, a geo entity and other dimensions like source and time period that were recorded with the original dataset. The data model is flexible such that each data point can be stored with a variable set of dimensions. The indicators appearing in the source dataset are mapped to the HHM indicator ontology so that data from various sources can be aggregated and combined.

The load process takes the curated and normalized values along with all its dimensions and associated metadata and loads it into an ElasticSearch data store. The ElasticSearch data store allows the flexibility to hold disaggregated value data along with all its dimensions, indexes them and is able to retrieve these values depending upon use cases and user requirements.

### Data access and aggregation service

A data access and aggregation service provides APIs with query parameters to access the ElasticSearch store and retrieve individual health indicators - geo entity values. These values are aggregated based upon the user queries and association rules; and provided to the visualization engine. This service is flexible and generic; and will evolve depending upon the use cases and dashboards designed on HHM.

### Web map service

The web map service is a web GIS stack consisting of a Postgres PostGIS datastore and a Geoserver tiling engine to produce raster, image and vector tiles. The web map service will curate and warehouse publicly available social - economic - environmental layers relevant for public health and infectious diseases. These include vegetation and land use layers, livestock distributions, meteorological layers, social and demographic layers, habitation layers as well as derived and proxy layers. The web map service will also build interfaces with other public and open geographical datasets that are growing like the World Environment Situation Room of the UNEP^24^, Hand-in-Hand data platform by Food and Agriculture Organization^25^, and the Resource Watch platform by World Resources Institute^26^.

### Visualization engine

The visualization engine is a front end component that will receive data in the form of JSON objects from the backend microservices and display it on the browser. The visualization engine will have a map based visualization, a tabular visualization and charts and plots based visualizations. The visualization engine will evolve and develop based on user needs. Future functionality will allow creation of dashboards and the ability to build, share and save custom visualizations.

## A beta version of the Health Heatmap

The Health Heatmap is conceived as a web application that will continuously evolve and change driven by user experience and demands. The technology design of the system must have the ability to be extensible and scalable; building features demanded by users and scalable as usership increases. A perpetual beta model of software design and deployment is ideally suited for the HHM platform in its initial stages^27^. The software architecture will allow frequent and continuous update of features and functions while maintaining the basic structure and data model.

The HHM is built as a set of microservices interacting with each other through web services. Each microservice is a compact functional unit with its own data and application logic. A semantic version system allows independent development and management of these microservices where the data store as well as application can independently evolve. Further, all these microservices are open source published on github which can be reused in other web applications and utilities. These microservices applications are shared with other web applications being developed by a collaborative team of the India Biodiversity Portal^28,29^.

Data has been curated and aggregated from NFHS - 3 & 4, AHS (2012-13), Health and Family Welfare Statistics 2017, and IDSP. There are more than 300,000 data points extracted from these sources across over 200 health indicators. Historic trends starting from the 1980s are available for some of the indicators while most of the data lie between 2010 and 2020. Geographic layers have been derived from census maps of India, land-use, land-cover layers, and meteorological layers.

The HHM provides the ability to navigate the data that has been curated and deployed depending upon the use cases and user requirements. We have deployed three use cases exposing the HHM data in the first version and have the ability to build more use cases as well as allow the user to cut-slide-dice the data depending upon user requirements, download the data for research analysis and action as well as build fresh public use cases on the platform.

### The following use cases have been built

#### An interactive visualization of the state of health across all districts of India

Provides a quick overview of the health status of each district through national rankings based on aggregation of selected indicators to create a composite index. A set of indicators for the district are scaled and normalized to allow aggregation and comparison. We have used a method inspired from the methodology used by NITI Ayog in creating the Aspirational Districts Program.^8^ This interactive atlas allows a person to select the set of indicators and visualize the composite index on our map interface.

#### An interactive visualization of infectious diseases

A page to visualize trends of infectious diseases across time and geography. Helps an individual gain perspective on the relative burden of infectious diseases across these dimensions. (See Figure 5)

**Figure 5:**
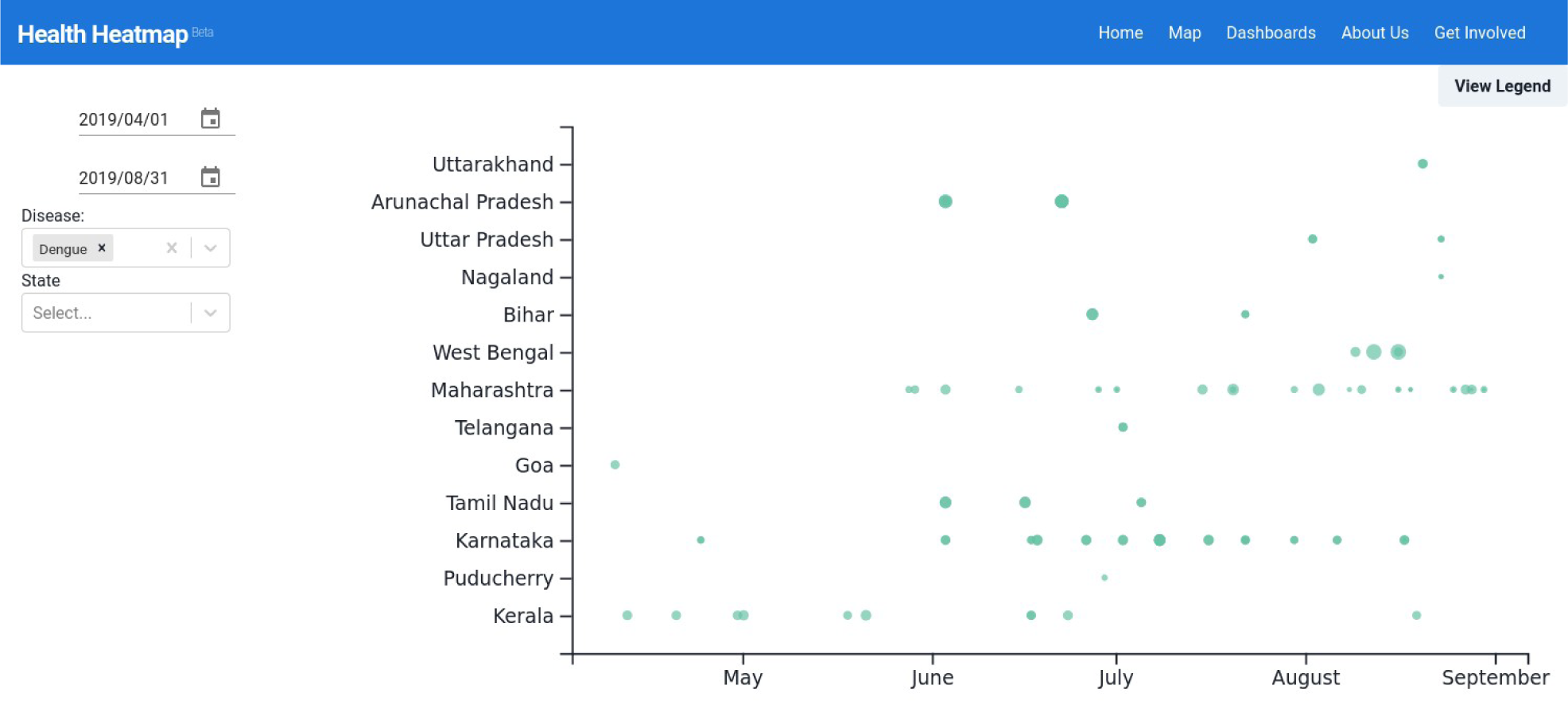
A screenshot of the page with interactive exploration of infectious diseases.

#### A deep dive exploration of the health data curated on the HHM platform

The complete HHM dataset visualized through all the map layers. Gives maximum control to the user on mixing and matching various indicators to arrive at their own interpretations. Filters that restrict dimensionality are also available. (See Figure 6)

**Figure 6:**
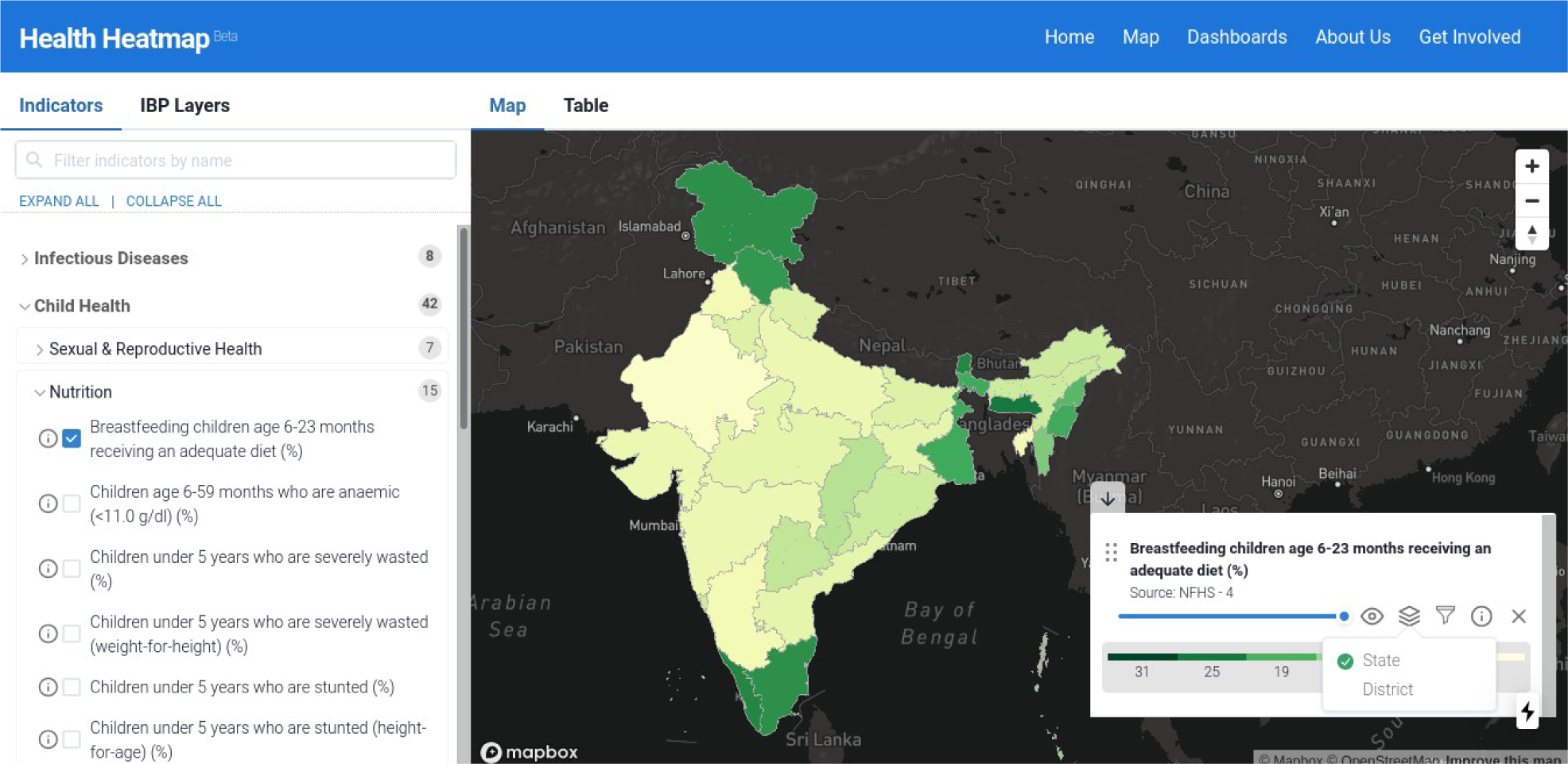
A screenshot of the deep-dive page. The panel for selection of indicators can be seen on the left.

The above are example use cases into the platform’s capabilities. Data having been digitized into an extensible model, the possibilities of developing specific use cases, rich visualizations. enabling user-driven explorations of data, public health monitoring and planning are opening up. We are actively seeking collaborations with universities, institutions, and individuals for expanding the scope and diversity of datasets available on the platform. We are developing software modules that enable more rigorous analyses such as the relationship between biodiversity and health, and external contributions are welcome on our public code repositories. Integration with R, an open source statistical computing environment, which would help advanced interactions and analysis of the data is also on the portal roadmap^30^. Further applications like predictive modeling are also on the horizon.

## Conclusion

A few years back, as noted by senior virologists in the country, India presented a poor picture of one health preparedness^31^. The governance structure and inter-sectoral coordination had been problematic, with human, animal and environmental health controlled by different ministries, with little cross-talk. Health policies had failed to even mention “zoonoses” and “emerging infectious diseases,” let alone break the silos or enable work in key areas.

In the course of building a beta version of the HHM platform, there were challenges of building a data model that would scale up and scale out. We have created an initial data model, but feedback from users would help in verifying and adapting the data model for scaling up. The public data available were distributed in different silos and needed to be considerably worked upon and curated. It would help if data were put out in more usable formats on the FAIR data principles. While we faithfully present the data with an attribution to the original source, the availability of data on a common platform allows comparisons and evaluation of data quality and reliability. Finally, HHM having been designed as a utility and service to society, the feedback from users and support from society would determine the future course of the platform.

This has been rapidly changing and while there are still miles to go, a semblance of evidence and digitally driven preparedness for healthcare seems to be emerging from the various national missions. The vision of an India that can leapfrog its healthcare system using innovation and scale in digital health technologies is the positive narrative that the nation is excited about. We believe that the creation of open platforms for warehousing and visualising health data of India as described here could be one of the pillars that the edifice of digital health can be built on.

The Health Heatmap of India (beta) portal can be accessed at https://healthheatmapindia.org

## Data Availability

All data is available through the portal itself at http://healthheatmapindia.org

## Acknowledgements and credits

The manuscript received valuable inputs, including a figure, from Prof Y Narahari at IISc. These are gratefully acknowledged. We also thank the independent reviewers for their valuable comments that helped improve the quality of the paper. We thank the software development team of Pradeep Thoma who designed and built all the user interfaces for the platform; and Piyush Kumar Singh and Prerana Singh for their active participation in the project.

This effort is part of the preparatory phase project of the National Mission on Biodiversity and Human Well-being, which is catalyzed and supported by the Office of the Principal Scientific Adviser to the Government of India.

## GLOSSARY NOTES

1. The *FAIR principles* - designed by academia, industry, funding agencies, publishers, and others - act as a guideline for those wishing to enhance the reusability of data, specifically by enhancing the ability of machines to automatically finding and using the data in addition to supporting reuse by individuals.
2. A *Heatmap* is a data visualization technique that makes it easy to understand complex data at a glance. In their widest sense, heatmaps include not just thematic geographical maps but also any kind of multidimensional data analysis that allows detection of clusters and prediction of hotspots.
3. AI/ML is an abbreviation that denotes Artificial Intelligence and Machine Learning which are the widely used methodologies for using modern computing engines and learning from data to improve the performance of systems and provide sophisticated predictive statistical models using highly non-linear regression techniques.Figure 1 is indicative of several applications that can be built on top of the foundation of a health heatmap. The gist of this paper describes an attempt to create the first box that instantiates how heatmaps provide valuable insights.

